# Delayed associations between air pollution and population health across the life course

**DOI:** 10.64898/2026.06.25.26356581

**Authors:** R. Alexander Bentley, Larisa Ozeryansky

## Abstract

Fine particulate air pollution (PM_2.5_) in the United States has fallen by roughly half since 2000, yet linked health outcomes such as diabetes and childhood ADHD have not improved in parallel. One reconciling possibility is that pollution exposure in early life produces health effects that emerge only years or decades later, after pollution itself has declined. Using two decades of U.S. county-level data, we relate annual PM_2.5_ estimates to birth outcomes, diabetes prevalence, and small-area estimates of childhood attention-deficit/hyperactivity disorder (ADHD) across short and long time scales. Within counties, changes in low birth weight rates are associated with changes in PM_2.5_ during the same year and the year prior to birth. At longer time scales, cross-county comparisons show that PM_2.5_ exposure is associated with higher prevalence of adult diabetes and ADHD after approximately a decade. Together, these patterns suggest that population-level health risks from air pollution may persist over decades, even as pollution itself declines.

## Introduction

In the United States, average fine particulate matter (PM_2.5_) concentrations have declined by nearly 50% since 2000 [1], yet many pollution-linked health outcomes have not improved in parallel [2, 3]. Air pollution remains a major global health risk [2], contributing to premature mortality through fine particulate matter [3–5] and to respiratory and cardiovascular disease [6–9]. The persistence of these health effects despite declining exposure suggests that the consequences of pollution may unfold over longer time scales than contemporaneous comparisons capture, with delays between early-life exposure and outcomes observed in childhood or adulthood [10].

Public health unfolds across multiple scales of time and space, from biomolecular processes to populations and from moments to generations [11]. Across development from the fetus through the stages of human life history, human biology and cognition respond to nutritional, environmental, and socioeconomic contexts [12–14]. While longitudinal studies at the individual level have long been central to public health research, large-scale environmental and population data have created new opportunities to study population health across longer time scales [15–17]. In the United States, examples include decades-long health consequences of sugar consumption [18, 19] and environmental pollution [2, 5]. Increasingly available county-level datasets now make it possible to examine lagged and multi-scale relationships between environmental exposures, life-course processes, and population health.

Pollution exposure early in life is detrimental not only to near-term health, but also to early childhood health and human capital outcomes later in life [10]. Adverse health effects may begin before birth. Because PM_2.5_ contains metals and reactive compounds capable of inducing oxidative stress, inflammation, and disruptions in placental function, prenatal exposure to environmental pollution has been linked to low birth weight [20, 21], epigenetic alterations in gene regulation [22], and impaired fetal growth and neurodevelopment [23]. Prenatal stress may further affect fetal development through elevated glucocorticoid signaling, which has been associated with a range of adverse outcomes after birth, including cardiovascular disease, psychiatric conditions, and altered neurodevelopment [24–26].

In early childhood, air pollution may increase neuroinflammation and disrupt the blood–brain barrier, facilitating the entry of airborne pollutants into the central nervous system [27]. Environmental exposures across prenatal and early-life stages may contribute to metabolic disruption, oxidative stress, and altered brain development [28–30]. Exposure to PM_2.5_ during these periods has been linked to cognitive deficits and neurodevelopmental disorders, including attention-deficit/hyperactivity disorder (ADHD) and autism spectrum disorder [31], which frequently co-occur [32, 33].

ADHD affects nearly 10% of children in the United States and represents a major population mental health concern with long-term consequences [34]. Air pollution during early developmental periods has been linked to elevated ADHD risk across diverse populations and exposure windows, with cohort and cross-sectional studies internationally reporting increased risk per unit PM_2.5_ exposure as well as effects on attention and working memory [35, 36]. Findings are not uniform, however, and some studies report null or attenuated associations at certain ages or for specific pollutants.

At the population level, both pollution exposure and pollution-linked health outcomes are geographically structured across the United States, mirroring the spatial distribution of socioeconomic disadvantage [37–43]. ADHD diagnosis rates, in particular, vary across demographic and socioeconomic groups in ways that closely track county-level income, education, and demographic composition [44–49]. This spatial coupling raises a question that contemporaneous regression cannot resolve: when pollution and socioeconomic disadvantage are entangled in space, do they also operate on the same time scale, or do they shape population health through different temporal pathways?

Most studies of air pollution compare environmental exposure and health outcomes measured over the same time period. This approach may overlook delayed associations when exposure occurs early in life but health outcomes emerge years later. The accumulation of two decades of nationwide environmental and health data now provides an opportunity to examine how associations between air pollution and population health vary across temporal scales.

The conceptual framework for this study (Figure 1) emphasizes the temporal dimension linking environmental exposures, prenatal development, and later health outcomes. Socioeconomic conditions affect both environmental exposures, such as air pollution, and the maternal environment during pregnancy, including stress and related factors. These influences operate through biological mechanisms such as placental function and fetal development, which may manifest in measurable birth outcomes, including low birth weight [51, 52], and contribute to later ADHD outcomes in childhood.

**Figure 1.**
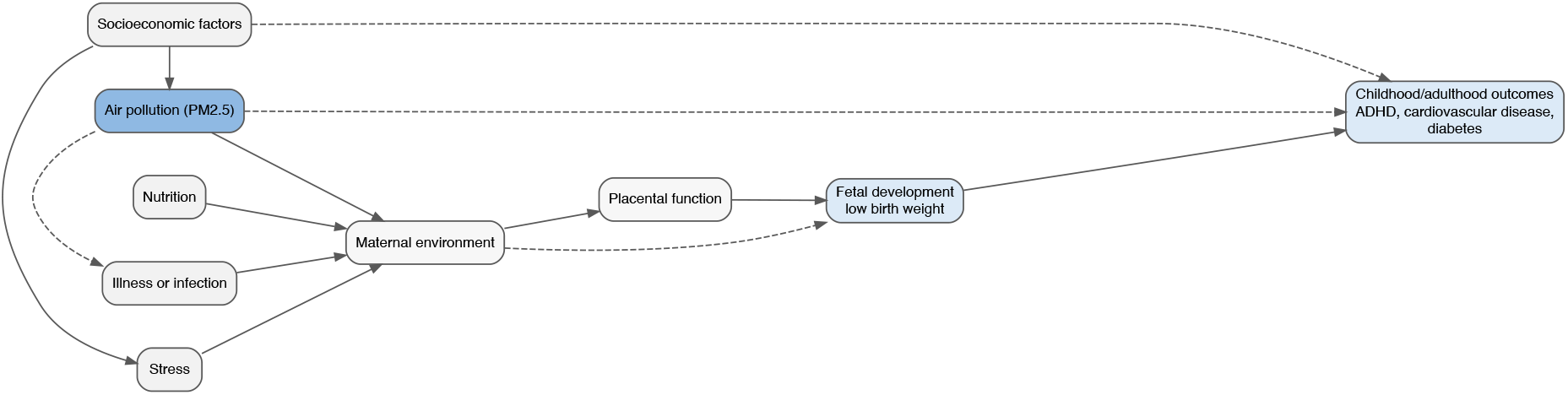
Conceptual framework linking environmental exposures, prenatal development, and later health outcomes. Blue boxes denote variables measured directly in population data, while gray boxes represent contextual or biological mechanisms. Dashed arrows indicate secondary pathways through which air pollution may influence infection risk or later health outcomes independently of birth weight. Adapted partially from [55]. The framework illustrates hypothesized pathways and is not directly estimated in the empirical analysis.

In this study, we examine population-scale associations between PM_2.5_ exposure and health outcomes in low birth weight, diabetes, and ADHD. Using U.S. county-level data, we test whether changes in pollution exposure within counties are associated with low birth weight, and whether persistently higher pollution levels are associated, years later, with higher ADHD prevalence. This highlights the importance of life-course timing in population health.

## Results

### County-scale variation and preliminary lagged correlations

Figure 2 shows how these county-scale variables pattern across the United States, including PM_2.5_, low birth weight, diabetes prevalence, and ADHD.

**Figure 2.**
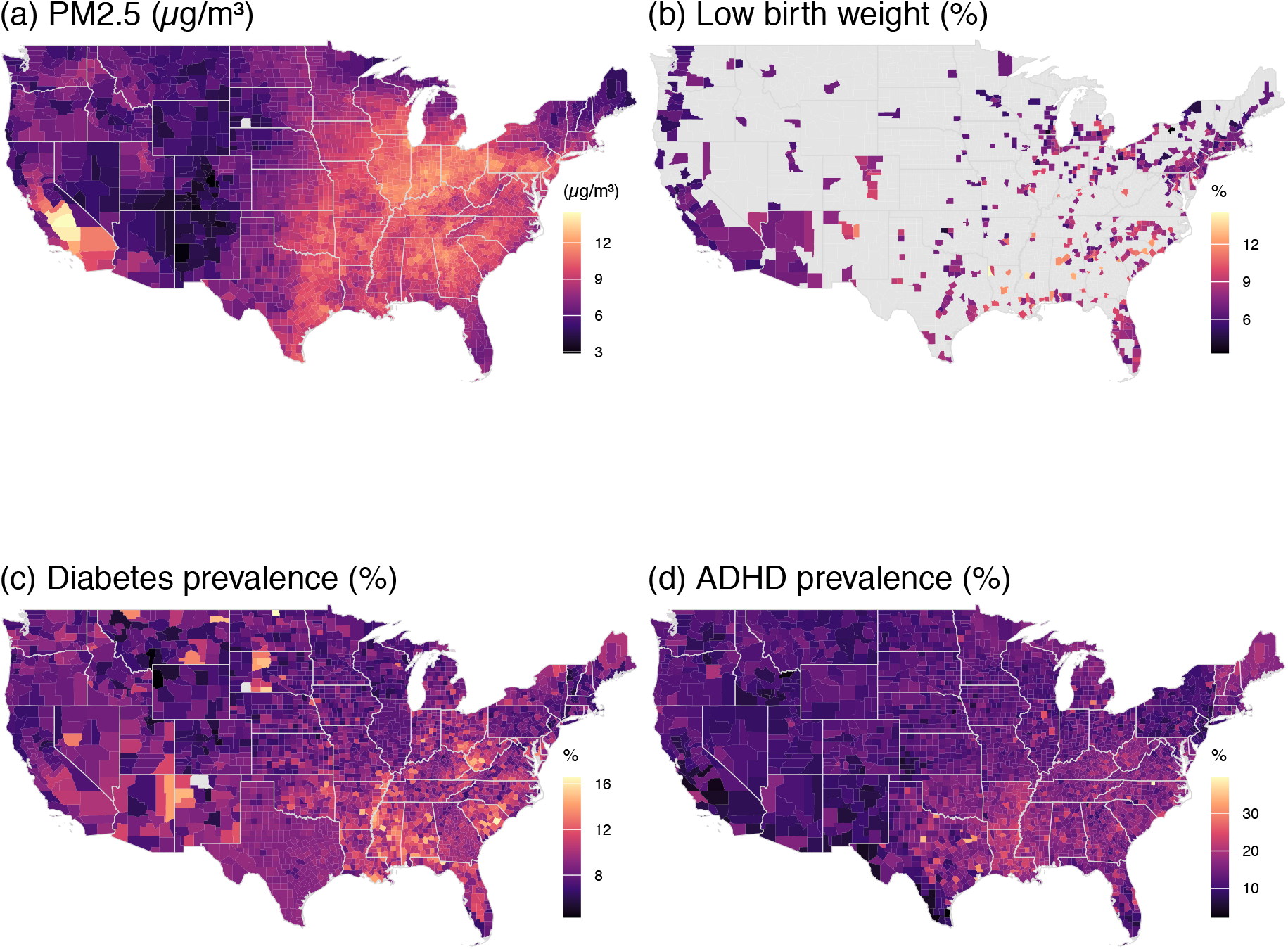
County-scale variation in air pollution and health outcomes in the United States. (a) Long-term average PM_2.5_ exposure, 2005–2015 [58, 59]. (b) Low birth weig4ht rate in 2016 among reporting counties [60]. (c) Diabetes prevalence in 2020. (d) ADHD prevalence among children aged 5–17 years, 2016–2018 [49].

Across the study period, the mean county-level low birth weight rate was approximately 8.2%, with values ranging from about 3.7% to 16.4% across counties and years (Figure 2b). As a preliminary descriptive check, we examined cross-county correlations between low birth weight rates in 2020 and lagged PM_2.5_ levels from the same counties. The contemporaneous correlation (lag 0) is small and not statistically significant (*r* = − 0.08), whereas correlations with earlier PM_2.5_ exposure (lag > 0) are positive and statistically significant, reaching *r* = 0.34 for PM_2.5_ levels ten years prior to birth (Figure 3a,b). In contrast, diabetes prevalence exhibits a more delayed association with PM_2.5_ (Figure 3c), with the strongest correlations occurring at lags of approximately 12–14 years (Figures 3c,d).

**Figure 3.**
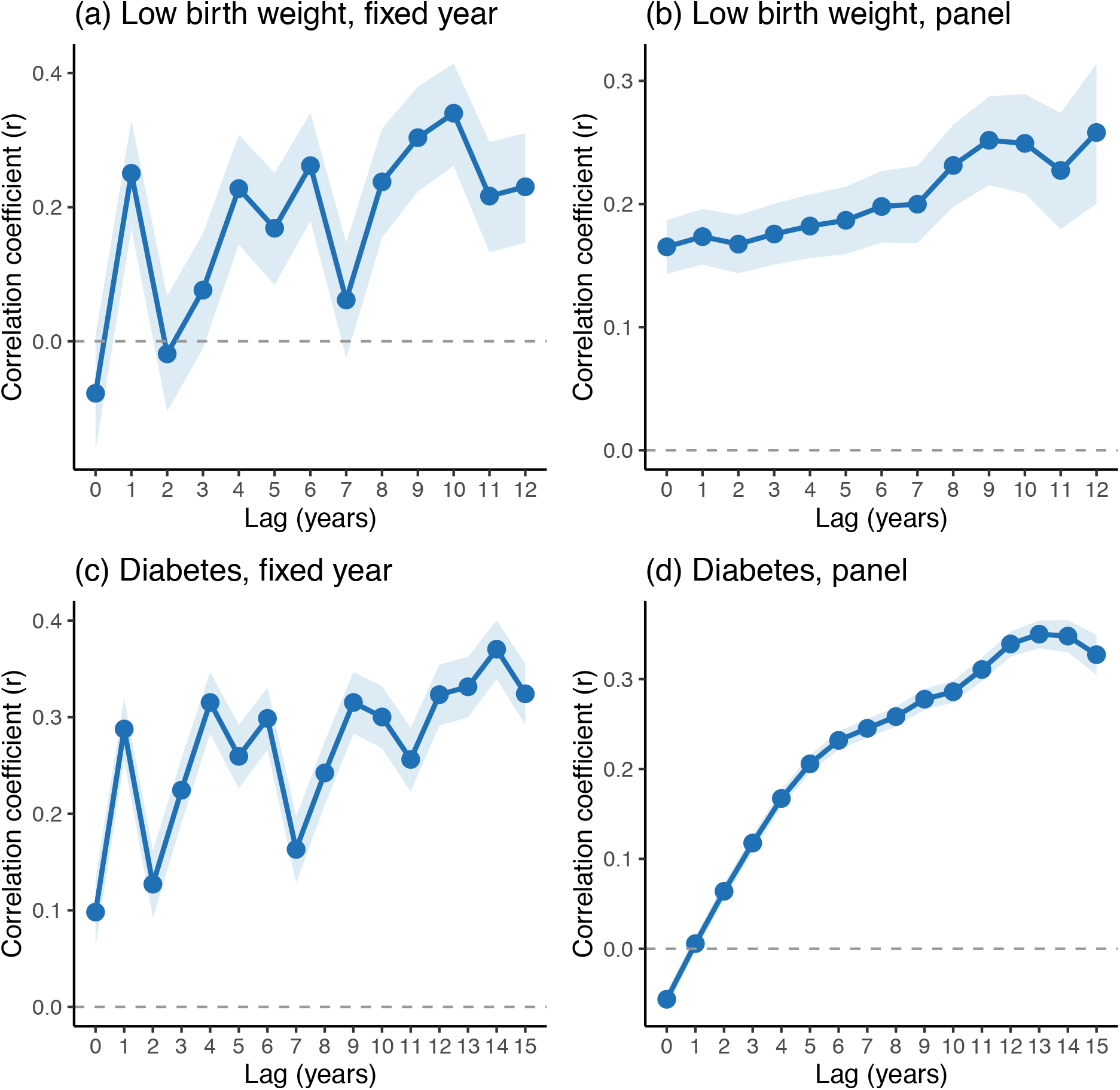
Lagged correlations between PM_2.5_ exposure and health outcomes. (a) Cross-county correlations between low birth weight in 2020 and PM_2.5_ lagged by 0–12 years in the same set of counties. (b) Panel correlations between low birth weight and PM_2.5_ lagged by 0–12 years using all available county–year observations. (c) Cross-county correlations between diabetes prevalence in 2020 and PM_2.5_ lagged by 0–15 years in the same set of counties. (d) Panel correlations between diabetes prevalence and PM_2.5_ lagged by 0–15 years using all available county–year observations. Shaded ribbons show 95% confidence intervals.

### Cross-sectional and within-county associations: low birth weight and diabetes

Table 1 summarizes regression results from the cross-sectional covariate model (eq. 2) and the county fixed-effects model (eq. 3) for both low birth weight and diabetes prevalence. Two patterns emerge that together motivate a multi-scale interpretation. First, in the cross-sectional covariate model (eq. 2), PM_2.5_ exposure is not significantly associated with low birth weight; rather, cross-county variation in low birth weight is explained primarily by socioeconomic structure — education, age, income, and economic connectedness — all of which are strongly significant (Table 1, left column). The persistent spatial geography of low birth weight across U.S. counties is, in this sense, a socioeconomic geography. Second, however, PM_2.5_ becomes a significant predictor once county fixed effects absorb these persistent differences (see below), pointing to a separate, shorter-time-scale role for pollution that the cross-sectional comparison cannot detect.

**Table 1.**
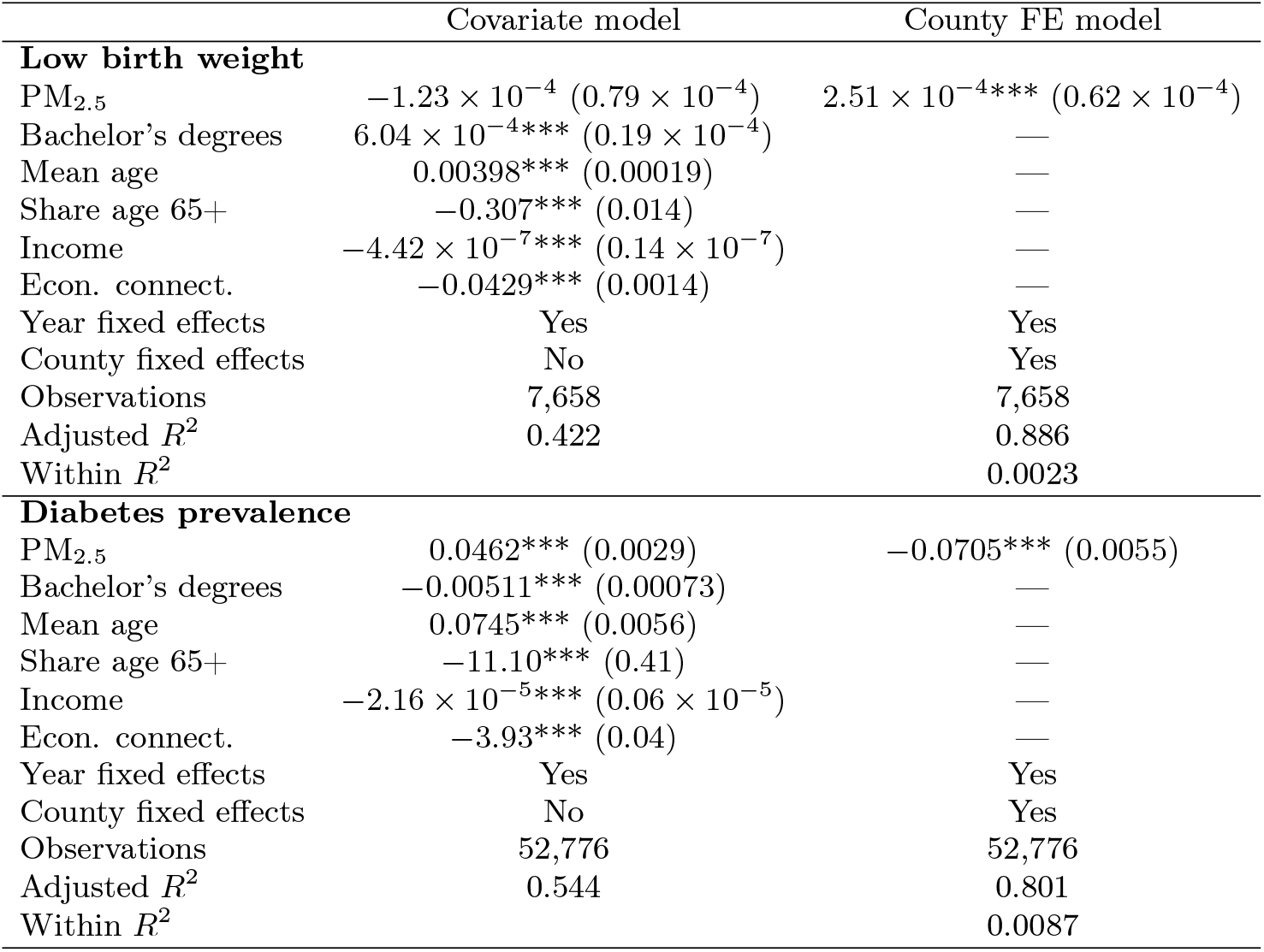
Regression estimates of associations between PM_2.5_ exposure and health outcomes. County-level covariates are included in column (1) and absorbed as fixed effects in column (2). Standard errors in parentheses. *** *p* < 0.001, ** *p* < 0.01, * *p* < 0.05. Low-birth-weight models include fewer observations due to omission of small-count counties.

In contrast, the same model for diabetes prevalence shows a positive and statistically significant association with PM_2.5_ exposure (Table 1, left column). Higher pollution levels are associated with higher diabetes prevalence across counties, although this cross-sectional relationship may reflect persistent spatial differences and broader temporal trends rather than within-county changes over time.

Estimating the county fixed-effects model (eq. 3) yields *n* = 7,658 county–year observations, with 576 county fixed effects and 14 year fixed effects (Table 1, right column). In this weighted regression, PM_2.5_ exposure is positively and significantly associated with low birth weight (Table 1, right column). The within-*R*^2^ of the model is modest (0.0023), indicating that PM_2.5_ explains only a small share of within-county changes in low birth weight over time. A one-unit increase in PM_2.5_ (1 µg/m^3^) within a county is associated with a 0.025 percentage-point increase in the low birth weight rate (SE = 0.000062, *p* < 0.001). For context, a 5 µg/m^3^ increase in PM_2.5_, about the range across U.S. counties (Figure 2a), is associated with an increase of about 0.13 percentage points in the low birth weight rate. Although modest in magnitude, such differences at the population level may affect a large number of births.

For diabetes prevalence, the county fixed-effects model (eq. 3) yields a negative and statistically significant association with PM_2.5_ (Table 1, right column). This negative within-county relationship contrasts with the positive cross-sectional association and reflects the opposing national trends of declining PM_2.5_ concentrations and rising diabetes prevalence over the study period.

### Lag structure of within-county associations

To examine whether earlier PM_2.5_ exposure predicts subsequent low birth weight within counties, we estimate lagged versions of the county fixed-effects model (eq. 3) in which PM_2.5_ is shifted backward in time (Figure 4a). The models include county and year fixed effects and are weighted by the total number of births in each county–year. The estimated coefficients are positive and statistically significant for lags of zero to three years but not for longer lags, consistent with short-term effects of pollution exposure during pregnancy.

**Figure 4.**
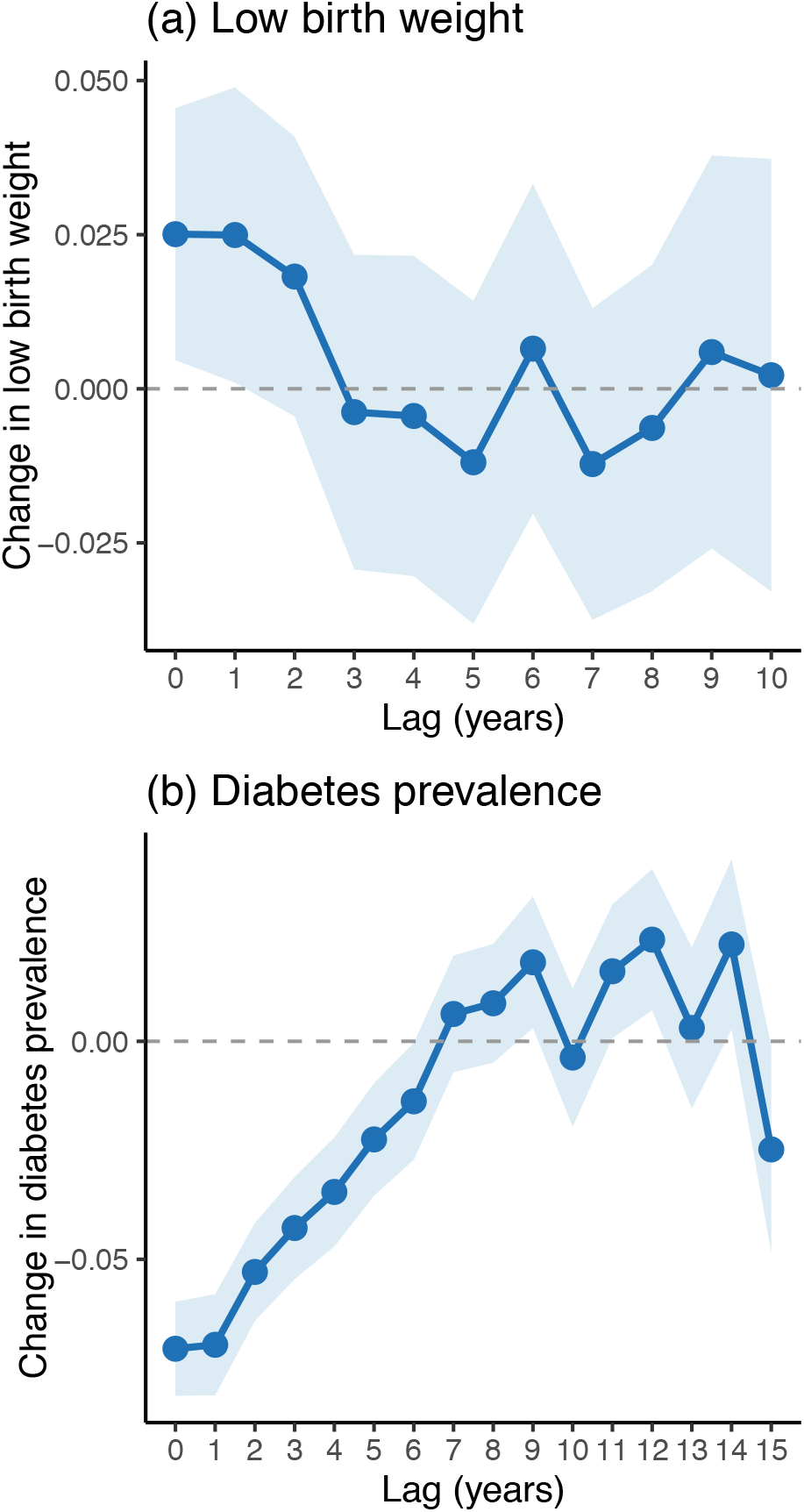
Within-county fixed-effects associations between PM_2.5_ and health outcomes. (a) Lagged association between county-level PM_2.5_ exposure and low birth weight (LBW). (b) Lagged association between PM_2.5_ exposure and diabetes prevalence. Panels show coefficients (with 95% C.I.) from county fixed-effects regressions relating outcomes to PM_2.5_ at lags of 0–10 years for LBW and 0–15 years for diabetes. LBW regressions are weighted by total births in each county–year.

In contrast, within-county fixed-effects estimates for diabetes prevalence show a different pattern. Because PM_2.5_ declined while diabetes prevalence increased over the study period, lagged PM_2.5_ is negatively associated with diabetes prevalence at short time lags (Figure 4b). At longer lags, the estimated coefficients become less negative and eventually positive after approximately seven years.

### Long-term PM_2.5_ exposure and childhood ADHD prevalence

We next examine whether long-term exposure to fine particulate matter is associated with county-level prevalence of ADHD among U.S. children. We estimate a sequence of cross-sectional regression models corresponding to eq. 4, relating ADHD prevalence to long-term PM_2.5_ exposure. The results, which progressively incorporate additional covariates, are shown in Table 2.

**Table 2.**
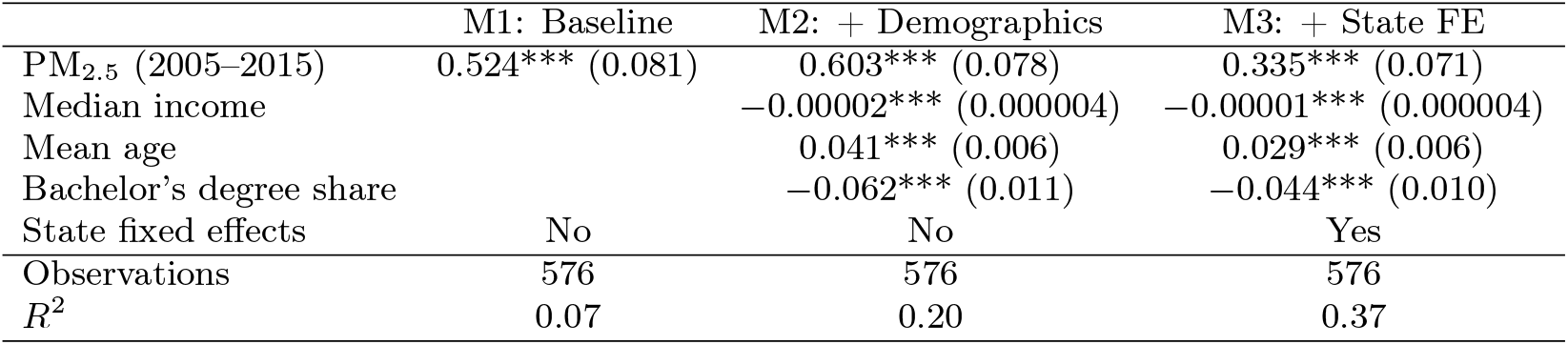
Long-term PM_2.5_ exposure and ADHD prevalence. Standard errors in parentheses. *** *p* < 0.001, ** *p* < 0.01, * *p* < 0.05.

|  | M1: Baseline | M2: + Demographics | M3: + State FE |
| --- | --- | --- | --- |
| PM<sub>2.5</sub> (2005–2015) | 0.524*** (0.081) | 0.603*** (0.078) | 0.335*** (0.071) |
| Median income |  | −0.00002*** (0.000004) | −0.00001*** (0.000004) |
| Mean age |  | 0.041*** (0.006) | 0.029*** (0.006) |
| Bachelor’s degree share |  | −0.062*** (0.011) | −0.044*** (0.010) |
| State fixed effects | No | No | Yes |
| Observations | 576 | 576 | 576 |
| $R^2$ | 0.07 | 0.20 | 0.37 |

In Model 1 (Table 2), the baseline specification, we estimate a cross-sectional regression of ADHD prevalence on long-term PM_2.5_ exposure. Counties with higher pollution levels exhibit significantly higher ADHD prevalence (*β* = 0.524 percentage points per *µ*g/m^3^ PM_2.5_, *p* < 0.00001), although the model explains a modest share of the variance (*R*^2^ = 0.07).

Model 2 (Table 2) adds county-level demographic covariates, including median income, mean population age, and the proportion of adults with a bachelor’s degree. Including these controls increases the estimated coefficient on PM_2.5_ (*β* = 0.603, *p <* 0.00001) and explanatory power (*R*^2^ = 0.20).

Because diagnostic practices and healthcare systems vary across states, Model 3 (Table 2) includes state fixed effects, thereby comparing counties within the same state. In this specification, the association between long-term PM_2.5_ exposure and ADHD prevalence remains positive and statistically significant (*β* = 0.335 percentage points per *µ*g/m^3^, *p* < 0.00001). Interpreting the coefficient magnitude, a difference of 5 *µ*g/m^3^ in long-term PM_2.5_ exposure corresponds to an estimated increase of approximately 1.7 percentage points in ADHD prevalence among children.

Figure 5a shows the partial regression relationship between county-level PM_2.5_ exposure (2005–2015 average) and ADHD prevalence after adjusting for income, mean age, educational attainment, and state fixed effects. Even after accounting for these factors, counties with higher long-term PM_2.5_ exposure exhibit higher ADHD prevalence among children. The partial regression slope (*β* ≈ 0.335 percentage points per *µ*g/m^3^) corresponds to the estimate from Model 3 in Table 2.

**Figure 5.**
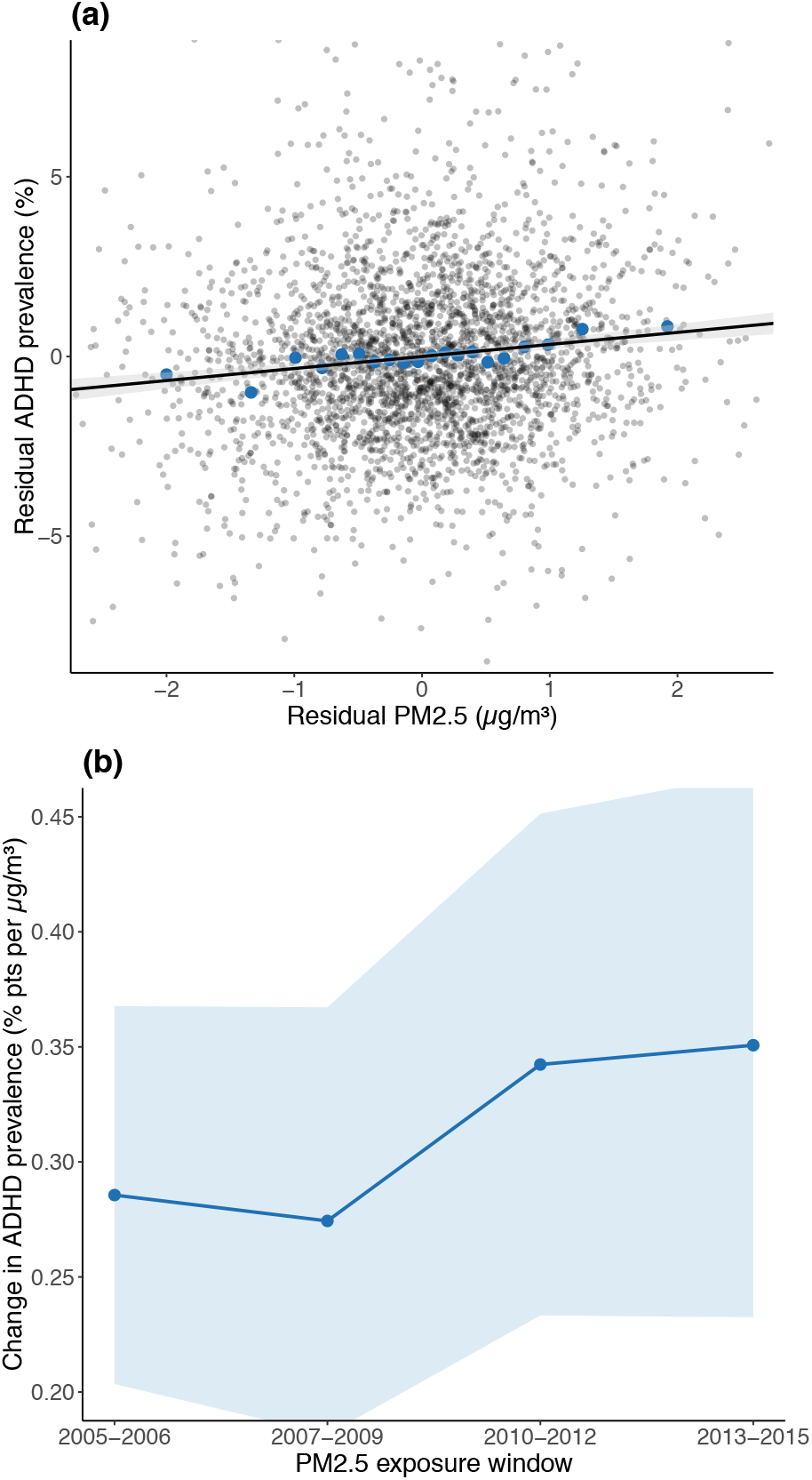
Long-term PM_2.5_ exposure and ADHD prevalence. (a) Partial regression plot showing the relationship between long-term PM_2.5_ exposure (2005–2015 average) and ADHD prevalence after adjusting for income, mean age, educational attainment, and state fixed effects. Gray points show individual counties, blue points show binned averages. The fitted linear relationship has slope *β* = 0.335. (b) Estimated associations (with 95% confidence) between county-level ADHD prevalence and PM_2.5_ exposure averaged over alternative exposure windows.

The results from Model 3 are robust to excluding counties in the top and bottom 1% of the PM_2.5_ exposure distribution (*β* = 0.360, *p* < 0.00001). Adding the economic connectedness index [53] as an additional covariate does not materially change the estimated association between PM_2.5_ exposure and ADHD prevalence, likely because it is highly correlated with other county-level characteristics already included in Model 3, including income, education, and age [53].

### Robustness and sensitivity checks

To assess whether changes in pollution are associated with changes in the composition of births, we estimate fixed-effects models using average maternal age as the outcome. Increases in PM_2.5_ are associated with slightly older maternal age within counties over time (*β* = 0.054 years per *µ*g/m^3^, *p* < 0.05), such that a 5 *µ*g/m^3^ increase in PM_2.5_ corresponds to an increase of approximately 0.27 years in average maternal age. However, the explanatory power of this relationship is very small (within *R*^2^ = 0.001). Compositional changes in maternal age are therefore unlikely to fully account for the observed associations between PM_2.5_ and birth outcomes. To the extent that older maternal age is associated with higher birth risks, this compositional change would, if anything, attenuate the observed relationship.

As a complementary check, we estimate the same fixed-effects specification using mean birth weight as a continuous outcome. The estimated association between PM_2.5_ and mean birth weight is negative but not statistically significant (*p* = 0.18), indicating that short-term within-county variation in pollution is not strongly associated with changes in average birth weight.

These results suggest that long-term PM_2.5_ exposure is associated with higher ADHD prevalence, even after adjusting for demographic factors and state-level differences in diagnostic practices. To assess sensitivity to the timing of exposure, we re-estimated Model 3—holding the covariates and state fixed effects constant—using alternative averaging periods for PM_2.5_ (2005–2006, 2007–2009, 2010–2012, and 2013–2015). Figure 5b shows that the estimated associations between PM_2.5_ and ADHD prevalence are consistently positive across these exposure windows, with coefficients of *β* = 0.286 for 2005–2006, *β* = 0.274 for 2007–2009, *β* = 0.342 for 2010–2012, and *β* = 0.351 for 2013–2015 (all *p* < 0.00001). The estimated effects increase modestly for exposure windows closer to the birth and early childhood years of the children in the sample, consistent with the possibility that early-life exposure to particulate pollution is particularly relevant for later ADHD diagnosis.

### Residual geographic patterns

Mapping the residual components of these models—after adjusting for socioeconomic variables and state fixed effects—helps illustrate the conceptual framework shown in Figure 1. The residual map of PM_2.5_ exposure (Figure 6a) shows pronounced geographic structure, likely reflecting topography (e.g., pollution trapping in California’s Central Valley), atmospheric transport patterns (e.g., contrasts between Appalachian valleys and the southeastern coast), and the spatial distribution of pollution sources such as power plants, industrial corridors, and petrochemical refining. This geographic structure becomes less pronounced in the map of residual diabetes prevalence (Figure 6c), which reflects longer time scales and additional intervening factors described in Figure 1. Even weaker spatial structure remains in the residual map of ADHD prevalence (Figure 6d).

**Figure 6.**
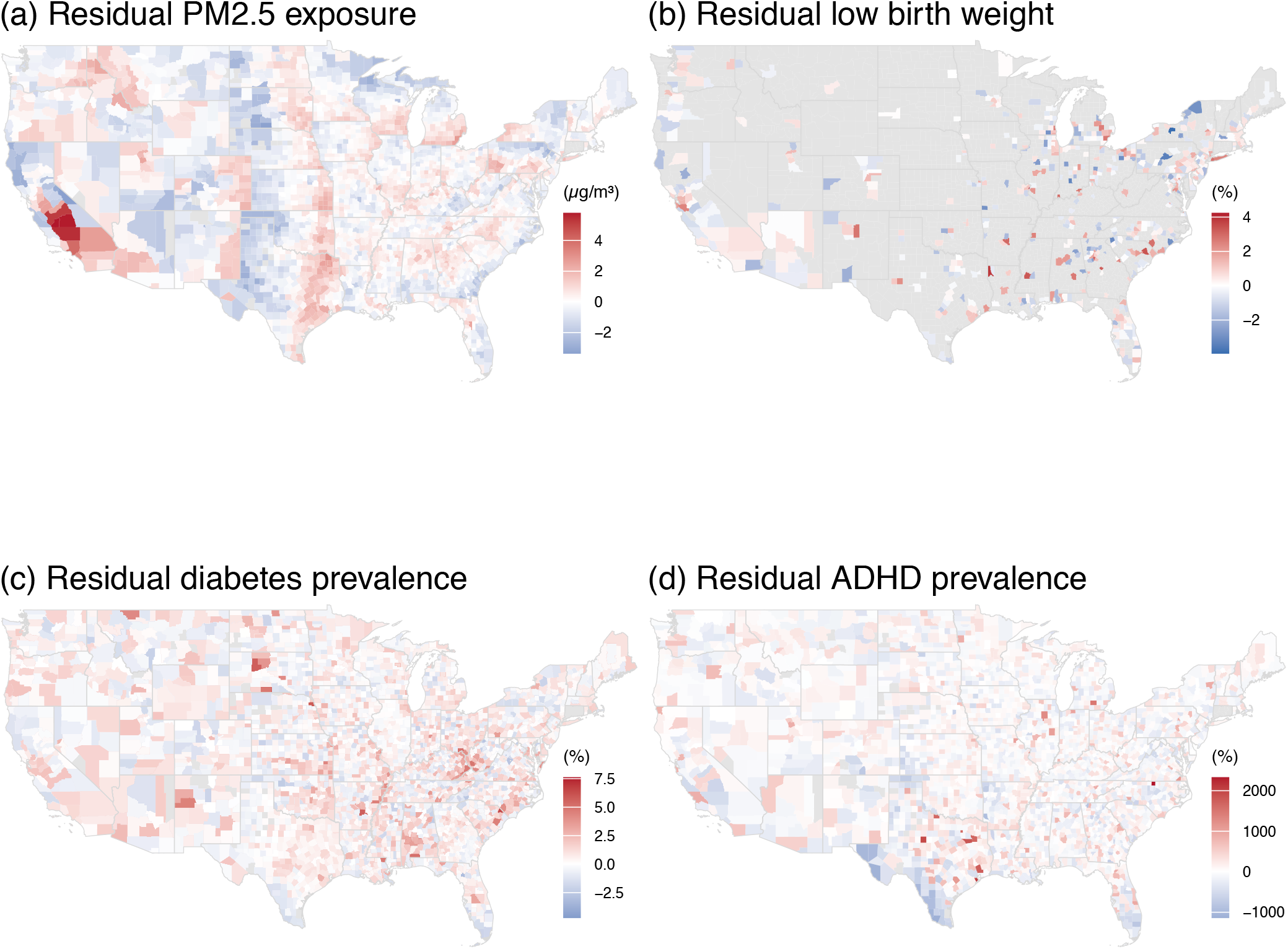
Residual county patterns after adjusting for socioeconomic covariates. Maps show residual values for (a) PM_2.5_ exposure, (b) low birth weight, (c) diabetes prevalence, and (d) ADHD prevalence after regression adjustment for income, mean age, educational attainment, and state fixed effects. Positive residuals indicate counties with higher values than predicted by the model, and negative residuals indicate lower-than-predicted values.

## Discussion

This analysis provides evidence of systematic, time-delayed associations linking fine particulate air pollution to a mental health outcome, ADHD, in childhood.

Using a county-level panel dataset, increases in PM_2.5_ within counties over time predict higher rates of low birth weight. The strongest associations occur for contemporaneous exposure and exposure one year prior to birth, consistent with epidemiological evidence that pollution exposure during pregnancy can affect fetal development. Because birth outcome data are only available for counties with reporting hospitals, the sample overrepresents more populous counties. As a result, the findings may not fully generalize to smaller or more rural counties, where both pollution exposure and health dynamics may differ.

The data also show associations at longer time lags—on the order of a decade—between long-term PM_2.5_ exposure and both diabetes and ADHD prevalence across U.S. counties. These relationships persist after adjusting for demographic and socioeconomic characteristics, including state-level differences in diagnostic practices.

The diabetes findings illustrate why temporal scale matters for interpretation. Within counties over the short term, PM_2.5_ is *negatively* associated with diabetes prevalence because the two trended in opposite directions during the study period: pollution fell while diabetes rose. Read as a contemporaneous effect, this would imply that pollution protects against diabetes, which is biologically implausible. The pattern instead reflects the fact that adult-onset diabetes accumulates over years of metabolic exposure, and short-term within-county fluctuations in PM_2.5_ cannot capture that build-up. Consistent with this, the within-county coefficient becomes less negative and turns positive at lags of roughly seven years and longer (Figure 4b), and the cross-county association between long-term PM_2.5_ and diabetes prevalence is positive (Table 1). The negative short-run within-county estimate is therefore not a contradiction of the cross-sectional pattern but a feature of the same underlying process viewed at the wrong time scale.

Because they are time-delayed through the life course, the health effects of air pollution may persist even as PM_2.5_ levels have declined overall in the United States [3, 5, 23, 50]. Counties with higher long-term PM_2.5_ exposure tend to exhibit higher ADHD prevalence among children approximately a decade later. Because the ADHD estimates represent children aged 5–17 years across multiple birth cohorts, the present analysis cannot distinguish precisely between prenatal and postnatal exposure mechanisms.

The spatial patterns of the residual maps also suggest this progression. The map of residual PM_2.5_ exposure (Figure 6a) retains a geographic imprint reflecting topography, atmospheric transport, and pollution source regions. This spatial structure becomes weaker for residual diabetes prevalence (Figure 6c) and weaker still for residual ADHD prevalence (Figure 6d). As envisaged in Figure 1, the effects of environmental exposures, as they interact with biological, behavioral, and social processes, become less direct over time.

Read together, these patterns suggest that pollution and socioeconomic disadvantage shape population health on different temporal scales. Socioeconomic structure — income, education, age composition, and economic connectedness — accounts for the persistent geography of pollution-linked health outcomes across U.S. counties, while pollution itself contributes to shorter-run within-county dynamics and to longer-run effects that emerge years after exposure. Both kinds of inequality therefore matter, and they matter through different pathways: SES sets the durable spatial landscape on which health outcomes are distributed, while pollution exposure is one mechanism through which that landscape is reproduced and amplified across the life course. Stress is a plausible linking pathway, disproportionately affecting lower-income populations [15, 54] and influencing birth outcomes through elevated cortisol levels linked to preterm labor and low birth weight [55–57]. Substantial racial, ethnic, and socioeconomic disparities in pollution exposure persist in the United States [3, 50], with county-level disparities in life expectancy [37, 43], indicating that the lagged health effects of pollution accumulate unevenly across the same populations who already bear higher exposure.

The associations reported here are descriptive rather than causal: the analysis does not exploit exogenous variation in air pollution, and estimates may reflect unobserved time-varying factors such as changes in local economic conditions, healthcare access, or public health policy. Several additional features of the data shape what can be inferred from them. The aggregated county-level design may mask within-county heterogeneity in exposure and health outcomes; the birth-outcomes panel is limited to counties with sufficient reporting, which skews toward more populous areas; multi-year exposure averages cannot precisely separate prenatal from postnatal windows for ADHD; and small-area estimates of ADHD and diabetes prevalence carry their own uncertainty, as does population mobility across counties over time. Using county-level data for the United States, this study examines how exposure to fine particulate air pollution relates to health outcomes across different stages of the life course. We find that increases in PM_2.5_ within counties over time are associated with higher rates of low birth weight, with the strongest associations occurring for exposure during pregnancy and the year prior to birth. At longer time scales, counties with persistently higher PM_2.5_ exposure tend to exhibit higher ADHD prevalence among children. These findings contribute to a growing literature linking air pollution to neurodevelopment by highlighting the temporal dimension of this relationship. Because exposure may occur prenatally while ADHD is typically diagnosed years later, understanding the timing of associations is essential. The results suggest that population-level patterns linking pollution and ADHD may unfold over time scales on the order of a decade.

The findings are consistent with existing evidence that air pollution may contribute to ADHD risk, and more broadly underscore the importance of incorporating temporal dynamics and cumulative exposure when investigating environmental influences on health across the life course.

## Methods and data sources

### PM_2.5_ exposure

Annual PM_2.5_ estimates were derived from satellite-based global gridded data combining aerosol optical depth retrievals, chemical transport modeling (GEOS-Chem), and ground-based observations [58, 59]. We used annual mean total PM_2.5_ concentrations at 0.01° × 0.01° resolution (NetCDF format), available from https://sites.wustl.edu/acag/surface-pm2-5/. Gridded estimates were aggregated to the county level using population-weighted averages to obtain annual mean PM_2.5_ concentrations for each U.S. county beginning in 2000.

### Birth outcomes

Birth outcome data were obtained from the CDC WONDER Natality database [60]. County-level counts of births by weight category were downloaded from the “Infant Birth Weight (12-category)” table for 2007–2024. A county-year measure of low birth weight was calculated as the number of births weighing under 2500 grams [61] divided by the total number of births in that county-year. After merging these data with the PM_2.5_ dataset by county FIPS code and year, the final analytic dataset contained 7,658 county–year observations spanning 2007–2020 across 576 counties. Because CDC WONDER suppresses data for counties with small numbers of births, low birth weight estimates are available only for counties meeting minimum reporting thresholds.

### Diabetes prevalence

Annual, age-adjusted estimates of diagnosed diabetes prevalence among adults aged 20 years or older were obtained from the U.S. Diabetes Surveillance System (USDSS), maintained by the Centers for Disease Control and Prevention [62]. The resulting dataset provides comparable estimates of diabetes prevalence across U.S. counties over time.

### ADHD prevalence

County-level ADHD prevalence estimates were obtained from [49], who generated small-area estimates using restricted data from the National Survey of Children’s Health (NSCH). Their analysis focused on approximately 70,000 children aged 5–17 years in the 2016–2018 survey waves. County-level prevalence estimates were produced using a small-area estimation framework combining survey responses with demographic and geographic predictors. The resulting dataset provides ADHD prevalence estimates and confidence intervals for U.S. counties, linked by county FIPS codes. National ADHD prevalence in the sample ranged from 11.5% to 14.4% (95% CI), with substantial geographic variation [49].

### Socioeconomic covariates

County-level covariates included median income, mean population age, and the proportion of adults with a bachelor’s degree, available from U.S. Census data (https://data.census.gov/table/ACSST1Y2021.S1501). From the 2018–2022 American Community Survey (ACS S1501) 5-year estimates on educational attainment, we used the percentage of adults aged 25 years and over with a bachelor’s degree or higher at the county level.

As an additional covariate, we incorporated an economic connectedness index [53], measured in May 2022 using social network data from approximately 70 million Facebook users aged 25–44. Economic connectedness is defined as the share of friends with above-median socioeconomic status among individuals with below-median socioeconomic status, providing a measure of cross-class social interaction at the county level [53].

### Analyses

We estimated linear regression models to examine the time-lagged relationship between annual PM_2.5_ exposure and county-level rates of low birth weight. The lag structure captures both short-term exposure windows relevant to prenatal and perinatal outcomes and longer-term accumulation processes that may influence chronic conditions such as diabetes. We estimated county fixed-effects models that include indicator variables for both counties and years:

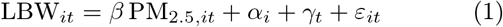

where *i* indexes counties and *t* indexes years. County fixed effects (*α*_*i*_) control for time-invariant characteristics of counties, while year fixed effects (*γ*_*t*_) control for nationwide trends affecting birth outcomes. Models were estimated using weighted least squares with weights equal to the number of births in each county–year. We estimated a series of lagged fixed-effects models in which PM_2.5_ was shifted backward in time by *k* years (*k* = 0 to 10), relating low birth weight in year *t* to PM_2.5_ exposure in year *t* −*k*.

As a baseline comparison, we also estimated contemporaneous cross-sectional regression models including county-level socioeconomic covariates:

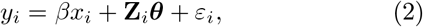

where *y*_*i*_ is the low birth weight rate in county *i, x*_*i*_ is PM_2.5_ exposure, and **Z**_*i*_ is a vector of county-level socioeconomic covariates (income, educational attainment, mean age, and economic connectedness) with corresponding coefficient vector ***θ***. Because these covariates primarily capture persistent differences between counties, we next estimated a model with county and year fixed effects:

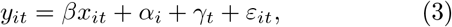

where *y*_*it*_ is the low birth weight rate in county *i* in year *t*, and *x*_*it*_ is PM_2.5_ exposure. In this specification, time-invariant county characteristics are absorbed by the county fixed effects.

We next estimated analogous models relating PM_2.5_ exposure to age-adjusted diabetes prevalence among adults aged 20 years or older. We estimated both covariate-adjusted cross-sectional models and county fixed-effects models, followed by lagged fixed-effects models in which PM_2.5_ was shifted backward by *k* years (*k* = 0 to 15), relating diabetes prevalence in year *t* to PM_2.5_ exposure in year *t* −*k*.

For analyses relating pollution exposure to ADHD prevalence, county-level PM_2.5_ averages were calculated for the period 2005–2015 to represent long-term exposure among children represented in the NSCH sample (ages 5–17 years in 2016–2018). Cross-sectional regression models related county-level ADHD prevalence to long-term PM_2.5_ exposure while adjusting for county socioeconomic characteristics and state fixed effects. Unlike the low-birth-weight and diabetes analyses, ADHD prevalence was available only as cross-sectional small-area estimates rather than a county–year panel. Consequently, lagged fixed-effects models could not be estimated, and temporal sensitivity was instead assessed by comparing alternative PM_2.5_exposure windows. The general specification is:

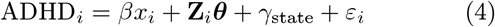

## Data availability

All data used in this study are publicly available. Annual PM_2.5_ estimates are available from the Atmospheric Composition Analysis Group (https://sites.wustl.edu/acag/surface-pm2-5/). Birth outcome data were obtained from the CDC WONDER Natality database. Diabetes prevalence estimates were obtained from the U.S. Diabetes Surveillance System. County-level ADHD prevalence estimates were obtained from Zgodic et al. [49]. County-level demographic covariates were obtained from the U.S. Census American Community Survey. The datasets generated and analyzed during the current study are available from the corresponding author upon reasonable request.

## Competing interests

The authors declare that they have no competing interests.

## Author contributions

R.A.B.: conceptualization, formal analysis, methodology, investigation, visualization, writing—original draft, review and editing; L.O.: writing— review and editing.

## Ethics and Consent to Participate

The study uses publicly available, aggregated county-level data from secondary sources and does not involve human participants, identifiable personal information, or intervention with individuals. Consequently, ethics committee approval and informed consent were not required.

## Funding

This research received no specific grant from any funding agency in the public, commercial, or not-for-profit sectors.

## Notes

### Competing Interest Statement

The authors have declared no competing interest.

